# Artificial Intelligence in Outpatient Primary Care: A Scoping Review on Applications, Challenges, and Future Directions

**DOI:** 10.1101/2025.05.12.25327223

**Authors:** Stacy Iannone, Amarpreet Kaur, Kevin B. Johnson

**Affiliations:** Division of Biomedical Informatics, Department of Biostatistics, Epidemiology, and Informatics, Perelman School of Medicine at the University of Pennsylvania, Philadelphia, Pennsylvania, United States; Clinical Futures Lab, Department of Pediatrics, Children’s Hospital of Philadelphia, Philadelphia, Pennsylvania, United States; David L. Cohen University Professor, Division of Biomedical Informatics, Department of Biostatistics, Epidemiology, and Informatics, Perelman School of Medicine at the University of Pennsylvania, Department of Pediatrics, Children’s Hospital of Philadelphia, Philadelphia, Pennsylvania, United States

**Keywords:** Scoping Review, Artificial Intelligence, Clinical Decision-Making, Outpatient Care, Health Information Systems, Patient Care Management, Primary Care

## Abstract

**Background:** Artificial intelligence (AI) has the potential to revolutionize clinical decision-making and significantly improve patient outcomes in outpatient primary care. AI technologies, including machine learning, deep learning, and transformers, enhance diagnostic accuracy, risk prediction, personalized treatment, workflow efficiency, clinical documentation, and continuous patient monitoring. However, despite rapid advancements, the extent of AI implementation in outpatient primary care remains unclear. This scoping review explores how AI functions, undergoes trials or integrates into non-urgent outpatient primary care settings.

**Methods:** This scoping review follows PRISMA Extension for Scoping Reviews (ScR) guidelines. We searched five databases, including published and gray literature, to identify studies published between January 1, 2019, and November 22, 2024, using AI and primary care-related terms. We used Covidence, a web-based systematic review tool, to screen titles, abstracts, and full texts of English-language manuscripts. We then extracted data and categorized studies by research phase and AI application in primary care.

**Results:** We screened 3,203 manuscripts and found 61 met the eligibility criteria. Most studies (26) focused on model development, while only eight reported clinical trial results. AI applications included provider support (5) and radiological disease diagnosis (1). Most studies examined clinical decision-making, disease diagnosis, and risk prediction, but none addressed provider cognitive support, workflow automation, or risk-adjusted paneling. Despite AI’s potential, real-world implementation remains limited.

**Conclusion:** AI in primary care remains in the developmental stage, with minimal real-world use beyond ambient scribing, clinical decision support, and workflow automation. Researchers must collaborate with professional societies and industry partners to accelerate adoption, expand clinical trials, enhance AI education for providers and patients, facilitate model deployment, and conduct periodic assessments of real-world AI adoption trends to guide future integration.

## Background

As the National Academy of Medicine noted, integrating advanced artificial intelligence (AI) technologies into healthcare may offer an unprecedented opportunity to transform patient care.^1,2^ The literature contains numerous examples showcasing the promise of AI, including systematic reviews to assess potential roles in clinical care and research.^3–6^ More recently, the release of AI tools powered by large language models (LLMs), such as ChatGPT, has led to a dramatic increase in tools that can augment the creation of medical information for specific purposes, such as drafting patient portal message responses, information summarization, and enabling ambient scribing, in outpatient care settings.^7–9^

Despite AI’s demonstrated potential in hospital settings, less is known about how AI is currently used in outpatient primary care clinics and ambulatory care settings or its broader impact on the healthcare system.^10,11^ Lin and colleagues note that mature AI systems may enhance primary care by impacting disease diagnosis, clinical decision-making, risk prediction, population risk-adjusted paneling, population management, providing medical advice to patients, and practice management. AI may also provide charting assistance by automating chart reviews and documentation, improving the integration of wearable device data into the decision-making process, and offering other forms of cognitive support to primary care providers.^12^ In light of these potential advancements, it is crucial to periodically assess the current state of AI implementation in primary care, identify existing research and deployment gaps, and encourage AI researchers to address these challenges.

## Objective

This scoping review examines the use of AI technologies in outpatient primary care and relevant ambulatory care settings. Specifically, it aims to summarize AI’s impact, identify gaps in the literature, and explore areas where AI holds the most significant potential to enhance primary care delivery.

### Materials and Methods

We conducted a scoping review using the Preferred Reporting Items for Systematic Reviews and Meta-Analyses (PRISMA) extension for Scoping Reviews (PRISMA-ScR) guidelines.^13^ We selected this methodology because it aligned with our objectives of describing the breadth of AI tools studied in primary care settings.

### Search Strategy and Selection Criteria

The research team began by conducting a comprehensive search across multiple databases, including PubMed, MEDLINE, Scopus, CINAHL, and Clinicaltrials.gov, for English-language studies published in peer-reviewed journals and research studies between January 01, 2019, and November 22, 2024. We developed our search strategy in collaboration with librarians at the University of Pennsylvania, Perelman School of Medicine, specializing in medical science and engineering. ur search strategy used a combination of key terms, subject headings, and standard abbreviations broadly related to AI, primary care, ambulatory care, and patient care. *Supplemental Appendix 1, Table A,* summarizes our final search strategy for each database used.

### Study Selection

Our review team (SLI, AK, KBJ) established inclusion and exclusion parameters prior to developing the search strategy and iteratively refined them to focus on the most relevant studies. We included peer-reviewed studies examining the use of AI/ML by healthcare providers in primary care settings.

We used Covidence, a web-based tool designed to streamline systematic reviews,^14^ to aid in screening and selecting unique articles based on the specified criteria. All three authors conducted the screening and selection process, ensuring they followed the PRISMA-ScR guidelines’ two-step approach. The team first screened articles based on titles and abstracts. To ensure consistency and to refine the inclusion and exclusion criteria, we conducted two trial runs, starting with 50 articles from PubMed, to align the reviewers’ understanding of the literature. Following consensus on the initial sample from PubMed, the team screened another 50 articles from Scopus to check for reduced inter-reviewer differences, which successfully diminished. Once alignment was achieved, the two reviewers proceeded to screen the remaining articles by title and abstract.

We excluded studies focused on inpatient care, emergency departments, nursing facilities, end-of-life or palliative care, and specialty conditions unlikely to be managed in primary care (e.g., neurology). Additionally, studies involved pharmacology, nursing, veterinary medicine, and non-healthcare sectors—e.g., environmental, agricultural, and social sciences. Data sources such as physician letters, book chapters, reviews, surveys, and focus groups were also excluded.

After removing duplicates, the three reviewers (SLI, AK, KBJ) independently screened article titles and abstracts for relevance using Covidence. Two authors evaluated each abstract reviewed against the eligibility criteria, with a third reviewer serving as a tiebreaker in cases of disagreement. We included full-text articles in the final analysis only if the two initial reviewers reached a concordant decision. In cases of discordance, all three reviewers reexamined the article and resolved the disagreement by consensus.

In the second step, two reviewers conducted a full-text review of the articles identified during the initial screening, carefully assessing all articles by applying the established inclusion and exclusion criteria to determine final eligibility.

### Data Extraction and Synthesis

A standardized data chart was developed to ensure consistency in data extraction across all included studies.

- Article citation details (authors, title, year)
- Country and targeted health organization (when applicable)
- Focus of innovation:

- Primary care provider Cognitive support
- Practice management
- Clinical decision-making
- Disease diagnosis
- Chart review/documentation
- Wearable integration
- Risk-adjusted paneling
- Medical advice
- Population health
- Risk prediction
- General discussion
- Type of study:

- Development
- Validation
- Protocol
- Trial
- Clinical use (descriptive or observational)

## RESULTS

Figure 1 summarizes the PRISMA-ScR screening and selection process. Our search strategy yielded 3,203 potentially relevant manuscripts from PubMed (*n*= 507), Scopus (*n*=285), CINAHL (*n*=2,004), and Clinicaltrials.gov (*n*=407). No additional references meeting our inclusion criteria were discovered through in-text citations.

**Figure 1.**
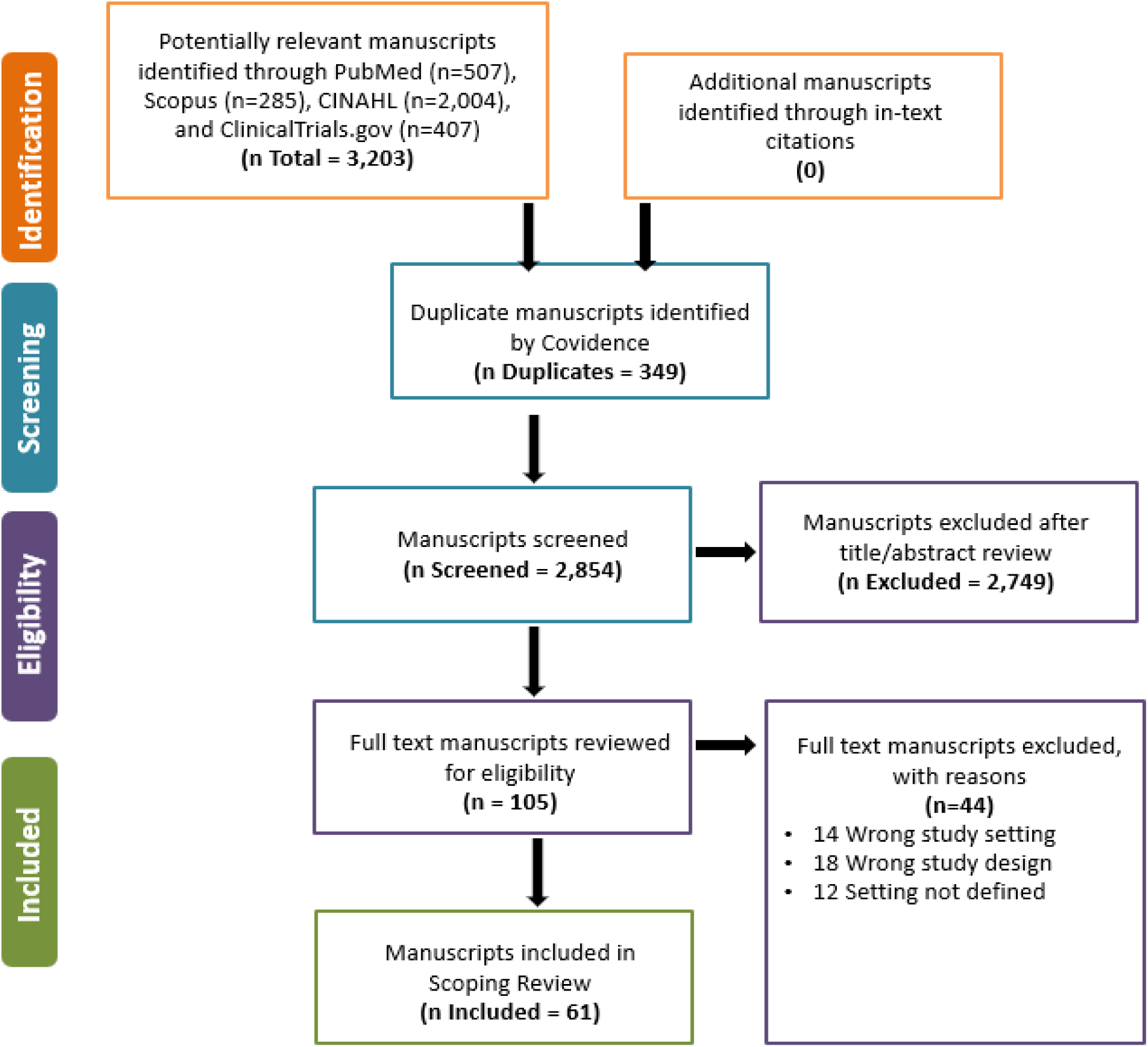
PRISMA flow diagram for a scoping review of eligible studies

After eliminating duplicates, 2,854 titles/abstracts were screened; 105 were deemed eligible for full-text review. Consensus was achieved for all disagreements concerning the inclusion of articles reviewed in their entirety. A total of 61 studies satisfied all criteria and were included in and summarized in the final analysis. A complete list of included studies is provided in *Supplemental Appendix A, Table C, Table D, and* Figure 1.

### Study Characteristics

*Appendix 1, Supplemental Figure A* provides an overview of the journals in which the included articles were published. The articles were published in various journals, including specialty journals focused on oncology, orthopedics, endocrinology, cardiology, and informatics.

*Appendix 1, Supplemental Table D* summarizes the key characteristics of the included publications, using a framework outlining the potential impact of AI on primary care.^12^ Manuscripts originated from investigators in the United States (n=35), Europe (n=8), and other countries (n=9), among these, 19% focused on family medicine, 35% on internal medicine, and 8% on pediatrics. Studies were conducted in various settings, including primary care within health systems, private practices, and other outpatient environments. A majority (n=26; 42%) of the studies were conducted across multiple locations. More than half (n=31; 59%) of the articles appeared in the literature before 2023, when GPT 3.0 was publicly available. ChatGPT, specifically, was publicly released on November 30, 2022.Six papers that suggested work in development were ideation rather than experimentation publications.^15–20^ A total of 26 (49%) of the included publications focused on AI models in early development.^17,18,21–45^ Additionally, 13 publications (25%) described model validation experiments.^46–60^ Eight articles (15%) described a planned or completed clinical trial in the areas of cardiology screening^61^, suicide prevention^62^, diabetes self-management education^63^, diabetic retinopathy screening^64^, automated insulin delivery^65^, emotional distress counseling^66^, fracture detection^67^, and hypertension self-care^68^. Trials varied in their design, ranging from non-randomized single-arm to randomized trials. Furthermore, five publications investigated the use of generative AI systems in clinical care^69–73^, and one publication described the use of an AI-based diagnostic system for COVID-19 pneumonia^74^.

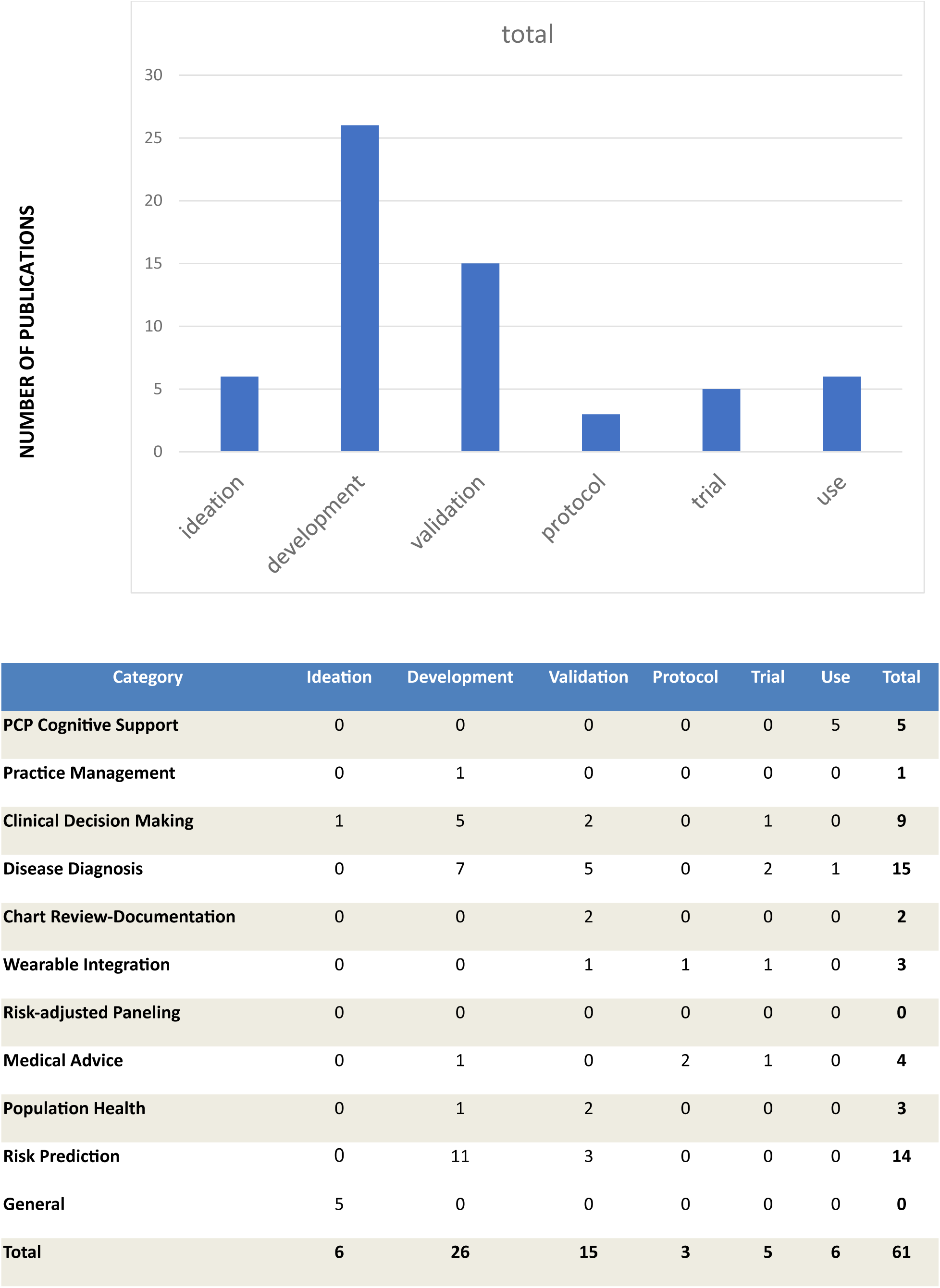

## DISCUSSION

This scoping review of AI applications in primary care revealed several challenges, highlighting both AI’s transformative potential and the complexities associated with integrating AI into primary care. While AI offers promising advancements across a broad range of primary care functions, it also exposes critical limitations in the current literature, such as variability in study quality (see Supplement) and the inherent heterogeneity of AI applications.

Despite the rapid proliferation of AI innovations, much of the work remains confined to the model development stage, with limited progress toward clinical implementation. Furthermore, many of these efforts are concentrated in some of the most complex areas of primary care, where implementation challenges are inherently higher. The scope of many studies remains narrow, with a significant number limited to single-site or small-scale projects, indicating that AI’s integration into real-world clinical practice is still in its infancy. Additionally, disparities in research focus areas and a limited number of large-scale planned or ongoing trials suggest a considerable delay in the equitable and widespread deployment of AI tools in primary care.

The decision to incorporate ideation papers in this review stemmed from the need to capture emerging perspectives, theoretical frameworks, and proposed innovations that had not yet undergone full clinical implementation. These papers provide valuable insights into the evolving direction of AI research and its anticipated role and impact in primary care. The interdisciplinary nature of AI in primary care is reflected in the diverse sources reviewed, spanning clinical research, health informatics, machine learning, and policy analysis. Given the complexity of AI integration, relying solely on clinical trials or implementation studies would have provided an incomplete picture. By considering a broad range of literature sources, this review presents a comprehensive understanding of AI’s trajectory, the gaps in research and implementation, and the systemic barriers that must be addressed to facilitate widespread adoption.

Several factors contribute to the gaps in AI research and implementation in primary care, including challenges in securing NIH funding for non-disease-specific projects, limited willingness among busy practices to test these emerging technologies, and the rapid deployment of commercial AI tools without academic partnerships or incentives for peer-reviewed validation. The concerns Kueper and colleagues. (2020) raised regarding the slow pace of AI integration remain highly relevant, underscoring the need for systemic change to overcome these barriers and advance AI’s responsible and effective deployment in primary care.

### Turning Challenges into Opportunities

A multipronged approach is needed to accelerate the responsible integration of AI into primary care. First, federal and state governments can play a pivotal role by funding large-scale clinical trials to assess AI applications’ safety, efficacy, and fairness. These trials could be led by learning health systems, as proposed by Johnson and colleagues^75^, or be sponsored by networks of care providers and professional societies, similar to initiatives led by the Pediatric Research in Outpatient Settings Network.^76^

Second, as LLMs such as ChatGPT become more prevalent, concerns around patient mistrust and the risk of AI models generating inaccurate outputs (hallucinations) must be addressed. Establishing rigorous standards for data security, privacy, and transparent reporting of LLM use is critical. Some communities have already proposed frameworks for responsible AI deployment, emphasizing accountability and transparent reporting of AI use in clinical settings.^2,78,79^

Third, AI developers often face challenges accessing primary care sites and data, hindering innovation and real-world testing. Strong partnerships between these AI developers and healthcare delivery experts will be essential. Collaborative efforts between healthcare providers, academic institutions, and technology companies could help bridge this gap while ensuring patient privacy and ethical data use. By fostering these interdisciplinary collaborations, ensuring equitable access to AI resources, and shifting focus toward large-scale implementation, AI can be effectively integrated into primary care, ultimately improving patient outcomes and healthcare delivery that aims to benefit all patients and providers.

#### Limitations

This review has several limitations stemming from its reliance on published literature, inherently excluding ongoing or unpublished studies. Consequently, it may provide an incomplete picture of the recent advancements or emerging trends in the field. Additional constraints include the following:

- **Language Restriction**: Only studies published in English were included, potentially overlooking relevant findings published in other languages.
- **Lack of Unified Definitions**: The absence of widely accepted standardized definitions for artificial intelligence (AI), machine learning (ML), and primary care introduces variability in study selection and interpretation.
- **Restricted Access**: The review did not incorporate insights from proprietary research, industry driven R&D efforts, or commercial developments, which may significantly influence AI advancements in primary care.
- **Limited Scope of Literature Queries**: Key repositories such as medRxiv, arXiv, and computer science conference proceedings were not queried. While these sources are unlikely to yield translational or clinical trial publications, their exclusion may still have omitted emerging technical innovations.

Moreover, the heterogeneous nature of AI applications across studies adds a layer of complexity. Variations in methodologies, study objectives, and evaluation criteria hinder the comparability of results and may affect the reliability and generalizability of this review’s findings. These disparities limit the ability to generalize conclusions across diverse healthcare contexts, thereby highlighting the need for caution in interpreting the review’s outcomes. One approach that could be done to alert the community to advances in this area is to leverage a period survey strategy focusing on the penetration of AI/ML tools in the primary care ecosystem. A similar strategy has begun to assess cognitive burden with some success.^80^

### Future Directions

Based on this review, future research must prioritize several key areas to advance the responsible integration of AI into primary care. A significant emphasis should be placed on conducting large-scale validation studies, publishing protocols, and disseminating successful AI-enabled tools and approaches to bridge the current gaps between development and clinical adoption. Researchers and information technology (IT) professionals play a vital role as they must address integration challenges to support pragmatic effectiveness trials, particularly those evaluating AI integration within real-world patient care settings. These integration challenges include A) technical barriers, such as ensuring compatibility with current electronic health records (EHRs) and maintaining robust data security and privacy concerns, and B) logistical and organizational concerns, including workflow adjustments, clinical staff training, and alignment with existing healthcare processes to promote seamless AI adoption.

Finally, researchers must explore the impact of AI on patient outcomes across diverse populations and healthcare settings. While AI technologies hold immense potential to transform healthcare by providing personalized and efficient care, their effects may vary significantly depending on patient demographics, underlying health conditions, and clinical environments.^81–84^ To ensure equitable patient access and benefits, researchers must investigate how AI impacts different groups, including various age ranges, socioeconomic backgrounds, and geographic locations. Future research prioritized by validation, integration, and inclusivity can drive meaningful advancements in healthcare delivery and patient outcomes, ensuring that AI-driven innovations are practical and broadly beneficial to the primary care ecosystem.

## CONCLUSION

This scoping review of 61tudies highlights the immaturity of AI and ML in primary care, with the predominant state of the research still focusing on development and validation rather than real-world implementation. To accelerate the maturation, testing, and development of AI and ML in primary care, clinical trials must be prioritized, coupled with the advancements of LLMs and the adoption of AI by commercial EHR vendors. These efforts will be critical in bridging the gap between AI innovations and its widespread adoption in clinical practices across diverse healthcare settings.

## Data Availability

All data produced in the present work are contained in the manuscript

## AUTHOR CONTRIBUTIONS

SLI, AK, KBJ conceptualized the scope of this review. AK and SI conducted the initial and full-text screenings. SI and KBJ drafted the manuscript with significant revisions and feedback from AK. We thank Lauren Malloy, Basam Alasaly, Rachel Wu, Kuk Jang, and Matthew Hill for reading and providing feedback.

## SUPPLEMENTARY MATERIAL

yes

## DATA AVAILABILITY STATEMENT

None

