## Appendices for "Artificial Intelligence in Outpatient Primary Care: A Scoping Review on Applications, Challenges, and Future Directions"

*Supplemental Table A: Summary of search terms and queries used in each academic literature database*

| Database | Concept | Search String | Operator |
| --- | --- | --- | --- |
| PubMed | Artificial Intelligence/ machine learning | ("Artificial Intelligence"[MeSH Terms] OR "AI"[Title/Abstract] OR "machine learning"[Title/Abstract] OR "deep learning"[Title/Abstract] OR "neural networks"[Title/Abstract] OR ambient[Title/Abstract]) | AND |
|  | primary/ ambulatory care | ("Outpatient Clinics, Hospital"[MeSH Terms] OR "outpatient"[Title/Abstract] OR "outpatient medicine"[Title/Abstract] OR "primary care"[Title/Abstract] OR "family medicine"[Title/Abstract] OR "ambulatory care"[Title/Abstract]) | AND |
|  | patient care | ("Patient Care"[MeSH Terms] OR "clinical encounter"[Title/Abstract] OR "patient encounter"[Title/Abstract] OR "healthcare improvement"[Title/Abstract] OR "patient care management"[Title/Abstract] OR "clinical decision making"[Title/Abstract]) | AND |
| Scopus | Artificial Intelligence/ machine learning | ( TITLE-ABS-KEY ( "Artificial Intelligence" ) OR TITLE-ABS-KEY ( "AI" ) OR TITLE-ABS-KEY ( "machine learning" ) OR TITLE-ABS-KEY ( "deep learning" ) OR TITLE-ABS-KEY ( "neural networks" ) ) | AND |
|  | primary/ ambulatory care | ( TITLE-ABS-KEY ( "Outpatient Clinics, Hospital" ) OR TITLE-ABS-KEY ( "outpatient" ) OR TITLE-ABS-KEY ( "outpatient medicine" ) OR TITLE-ABS-KEY ( "primary care" ) OR TITLE-ABS-KEY ( "family medicine" ) OR TITLE-ABS-KEY ( "ambulatory care" ) ) | AND |
|  | patient care | ( TITLE-ABS-KEY ( "Patient Care" ) OR TITLE-ABS-KEY ( "clinical encounter" ) OR TITLE-ABS-KEY ( "patient encounter" ) OR TITLE-ABS-KEY ( "healthcare improvement" ) OR TITLE-ABS-KEY ( "patient care management" ) OR TITLE-ABS-KEY ( "clinical decision making" ) ) | AND |
| CINAHL | Artificial Intelligence/ machine learning | (TX "Artificial Intelligence" OR TX "AI" OR TX "machine learning" OR TX "deep learning" OR TX "neural networks") | AND |
|  | primary/ ambulatory care | (TX "Outpatient Clinics, Hospital" OR TX "outpatient" OR TX "outpatient medicine" OR TX "primary care" OR TX "family medicine" OR TX "ambulatory care") | AND |
|  | patient care | (TX "Patient Care" OR TX "clinical encounter" OR TX "patient encounter" OR TX "healthcare improvement" OR TX "patient care management" OR TX "clinical decision making") | AND |
| ClinicalTrials.gov | Artificial Intelligence/ machine learning | ("Artificial Intelligence" OR "AI" OR "machine learning" OR "deep learning" OR "neural networks") | AND |
|  | primary/ ambulatory care | (“Outpatient Clinics, Hospital" OR "outpatient" OR "outpatient medicine" OR "primary care" OR "family medicine" OR "ambulatory care") | AND |
|  | patient care | (“"Patient Care" OR "clinical encounter" OR "patient encounter" OR "healthcare improvement" OR "patient care management" OR "clinical decision making") | AND |

*Supplemental Table B: Inclusion and exclusion criteria*

| **Category** | **Inclusion Criteria** | **Exclusion Criteria** |
| --- | --- | --- |
| Document Type | Peer-reviewed publications  Primary studies  Research articles | Physician letters or notes  Book or book chapter  Reviews (ex: systematic reviews, scoping reviews, etc.)  Surveys |
| Population | Ambulatory setting  Outpatient setting  Primary care  Mental health care in ambulatory setting  Rehabilitative care in ambulatory setting  EHR analysis within ambulatory clinical care (using EHR with AI) | Inpatient setting  Emergency department  Nursing care facilities  End of life care  Palliative care  Specialty care or condition (ex: COPD, neurology, etc.)  EHR analysis external to ambulatory care settings  Focus groups |
| Subject Area | Medicine  Clinical Informatics/ Engineering  Neuroscience  Psychology  Dentistry | Non-healthcare related fields (ex: environmental sciences, agricultural sciences, social sciences, etc.)  Nursing  Pharmacology & Pharmaceutics  Veterinary  Health Education  Healthcare management, finance or policies |
| Intervention/Exposure | Artificial intelligence  Deep learning  Machine learning  Neural networks |  |
| Outcome | Patient care enhancement/ satisfaction  Provider enhancement/ satisfaction | Patient hospitalization (context specific)  Performance assessment of healthcare personnel or programs  Provider/ Patient perspectives or interviews |

*Supplemental Table C: Characteristics of Included Publications*

| Attribute | References |
| --- | --- |
| Publication year | 2019, 2020, 2021, 2022, 2023-2024 |
| Study location | United states, Europe, Asia, Canada |
| Visit work | History review, symptom assessment, diagnosing, predicting, testing, treatment, education, follow-up planning, documentation |
| Technology used | AI-Based Models*, Machine Learning (ML) Models, Deep Learning (DL) Models, Natural Language Processing (NLP), Data and Sensor Technology, Mobile Health (mHealth), Clinical Decision Support Systems (CDSS), Wearable Devices, Predictive Models, Reinforcement Learning, Hybrid Neural Architecture, Generative AI  *AI-based models, Machine Learning (ML) Models, and Deep Learning (DL) Models are often used together in the same study, leading to multiple counts for a single study. Similarly, Data and Sensor Technology might be combined with Wearable Devices or Clinical Decision-Support Systems (CDSS), contributing to overlapping categories. |
| Study design | Quantitative (n=40), Mixed Methods (n=6), Qualitative (n=1), Protocol (n=1), Review (n=1), Case Study (n=1), Systematic Review (n=2) |
| Innovation | Decision algorithm development, Multicenter validation, Hybrid neural architecture, ECG-based machine learning, AI influence analysis, Natural language processing, Big data analytics, AI implementation in care, Deep learning support, mHealth and AI, Deep learning assistance, Machine learning models, Model validation, AI-based education, EHR data analysis, Study population enrichment, Wearable devices integration, Model application development, AI-driven toolkits, ChatGPT comparison, Screening tool implementation, Clinical trial, Tool validation, Risk prediction model, Performance comparison, Data-driven models, Prediction models, Deep learning validation, Multi-method validation, Symptom extraction, AI counselor, Nonrandomized trial, Diagnostic accuracy, Prediction model validation, Machine learning treatment, Clinical notes approach, Recurrent neural network, Reinforcement learning, Screening pilot, Multivariable models, image-based diagnosis |

Supplemental Figure A Sources of Included Articles

Sources of Included Articles

|  | Count |
| --- | --- |
| Academic Medicine | 1 |
| Addiction | 1 |
| Annals of Oncology | 1 |
| Annals of Family Medicine | 1 |
| Applied Clin Inform | 1 |
| BMC Medical Informatics and Decision Making | 2 |
| BMC Musculoskeletal Disorders | 2 |
| BMC Psychiatry | 1 |
| BMC Public Health | 1 |
| BMJ Open | 3 |
| Breast (Edinburgh Scotland) | 1 |
| Cancer epidemiology, biomarkers and prevention | 1 |
| Circulation | 1 |
| Clinical Orthopaedics and Related Research | 1 |
| Computational and Mathematical Methods in Medicine | 1 |
| Contemporary Clin Trials | 1 |
| Developmental Medicine and Child Neurology | 1 |
| Diabetes Care | 1 |
| Drugs | 1 |
| Fam Pract | 1 |
| Family medicine and Community Health | 2 |
| Frontiers in Neurology | 2 |
| Health care management science | 1 |
| IEEE | 1 |
| Inflammatory Bowel Diseases | 1 |
| International Jnl Med Inf | 1 |
| International journal of environmental research and public health | 1 |
| International Journal of Nursing Studies | 1 |
| Investigative Ophthalmology & Visual Science | 1 |
| JAHA | 1 |
| JAMA | 1 |
| JAMA Netw Open | 1 |
| JAMIA | 1 |
| JMIR | 3 |
| JMIR Cardio | 1 |
| JMIR Formative Research | 2 |
| JMIR Medical Informatics | 2 |
| Jnl of Electrocardiology | 1 |
| J Am Coll emerg Physicians Open | 1 |
| Mayo Clinic Proceedings | 1 |
| medRxiv | 1 |
| Med Inform Decis Mak | 1 |
| Nature Medicine | 2 |
| NEJM. AI | 1 |
| PloS One | 1 |
| Postgraduate Medicine | 1 |
| Respiratory Medicine | 1 |
| Sci Rep | 1 |
| Science Translational Medicine | 1 |
| The Lancet Digital Health | 1 |
